# Cost-Utility Analysis of PCSK9 Inhibitors and Quality of Life: A Two-Year Multicenter Non-Randomized Study

**DOI:** 10.1101/2023.12.05.23299557

**Authors:** José Seijas-Amigo, M. José Mauriz-Montero, Pedro Suarez-Artime, Mónica Gayoso-Rey, Francisco Reyes-Santías, Ana Estany-Gestal, Antonia Casas-Martínez, Lara González-Freire, Ana Rodriguez-Vazquez, Natalia Pérez-Rodriguez, Laura Villaverde-Piñeiro, Concepción Castro-Rubinos, Esther Espino-Faisán, Octavio Cordova-Arevalo, Moisés Rodríguez-Mañero, Alberto Cordero, José R. González-Juanatey, e Investigadores MEMOGAL

## Abstract

**Background:** The use of PCSK9 inhibitors in the management of cardiovascular disease has gained increasing attention. However, there is limited evidence on their cost-utility and impact on quality of life in real-world settings. This study aimed to conduct a real-world cost-utility analysis of PCSK9 inhibitors and evaluating their effects on quality of life.

**Methods:** A multicenter prospective study was conducted across 12 Spanish hospitals from May 2020 to April 2022, involving 158 patients. QoL was assessed using the EQ-5D questionnaire. Cost-utility analysis assessed the economic impact of PCSK9 inhibitors when used with standard care compared to standard care alone, calculating the incremental cost-effectiveness ratio (ICER).

**Results:** The study encompassed 158 patients with an average age of 61, predominantly male (66.5%). For patients initiating PCSK9 inhibitors, the cost of PCSK9 inhibitor treatment was €13,633.39, while standard therapy cost €3,638.25 over two years. Qualys for PCSK9 inhibitors stood at 1.648851948 over two years, compared to 1.454807792 for standard therapy. The results revealed favorable cost-utility outcomes, with an ICER of €51,427.72. Significant improvements were observed in the domains of mobility, self-care, daily activities, pain/discomfort, and anxiety/depression (p<0.001).

**Conclusions:** This study represents the first real-world cost-utility analyses of PCSK9 inhibitors. The findings support the economic justification and potential benefits of incorporating PCSK9 inhibitors into clinical practice. Healthcare decision-makers can consider these results when making informed decisions and reimbursement regarding the use of PCSK9 inhibitors in clinical practice.

Trial Registration clinicaltrials.gov **Identifier:** NCT04319081

## INTRODUCTION

Cardiovascular diseases (CVDs) remain a leading cause of morbidity and mortality worldwide, imposing a substantial economic burden on healthcare systems (1). Elevated low-density lipoprotein cholesterol (LDL-C) levels have been identified as a major risk factor for CVD development and progression. In recent years, the advent of novel therapeutic agents, such as proprotein convertase subtilisin/kexin type 9 (PCSK9) inhibitors, has provided promising avenues for effectively managing dyslipidemia (2).

PCSK9 inhibitors, such as evolocumab and alirocumab, have demonstrated remarkable LDL-C-lowering efficacy in randomized controlled trials (RCTs), leading to their approval and integration into clinical practice guidelines (3–4). However, the economic implications of these new therapies, especially in real-world settings, have garnered significant attention. Real-world studies provide crucial insights into the cost-effectiveness and value of interventions when implemented in routine clinical practice (5).

While cost-effectiveness analyses are commonly employed to assess the economic impact of healthcare interventions, the evaluation of cost-utility, which takes into account costs and health-related quality of life (QoL) outcomes, holds particular relevance. Cost-utility analysis utilizes preference-based measures, such as the EuroQol-5 Dimension (EQ-5D) questionnaire, to quantify changes in QoL and subsequently inform decision-making processes. In this context, it is important to emphasize that both cost-effectiveness analysis (CEA) and cost-utility analysis (CUA) provide valuable insights into the economic impact of healthcare interventions and contribute to informed decision-making. While CUA specifically focuses on health-related quality of life outcomes and incorporates QALYs, CEA assesses the overall cost-effectiveness of interventions. These complementary approaches collectively aid in assessing the value of healthcare interventions (6). Despite the increasing utilization of PCSK9 inhibitors, there remains a dearth of cost-utility evidence, especially derived from real-world studies. RCTs, although essential for establishing clinical efficacy, often lack generalizability to real-world populations and may not capture the broader economic impact of interventions (7).

Currently, there is a lack of cost-utility data derived from real-world studies for PCSK9 inhibitors, as quality of life and cost-utility variables are typically assessed primarily within the context of clinical trials. However, comprehending the economic impact and utility of these inhibitors in everyday clinical practice is vital for making well-informed choices (8).

The present study aims to investigate the cost-utility and quality of life outcomes of PCSK9 inhibitors using prospectively collected data from 158 patients over a 24-month follow-up period. The primary objective of this study is to assess the cost-utility of PCSK9 inhibitors, while the secondary objective was to evaluate changes in quality of life measured through the different domains of the EQ-5D questionnaire. By examining these comprehensive measures, this study aims to provide valuable insights into the economic impact and utility of PCSK9 inhibitors in real-world clinical practice, building upon the findings from the MEMOGAL study (ClinicalTrials.gov Identifier: NCT04319081).

## METHODS

### Study design

The MEMOGAL STUDY (NCT04319081) (9) is a multicenter, prospective study conducted in 12 Spanish hospitals with a double-arm, phase IV, open-label design. The study enrolled patients with familial hypercholesterolemia (FH) or established cardiovascular disease (CVD) who were initiating PCSK9i treatment for the first time with a follow-up of 2 years. The protocol received approval from the ethics committee and the Spanish Agency for Medicines and Health Products. Figure 1 illustrates the study design and patient disposition. In accordance with the Consolidated Health Economic Evaluation Reporting Standards (CHEERS) guidelines (ref), this study has been conducted and reported to ensure comprehensive and transparent reporting of health economic evaluations.

**Figure 1.**
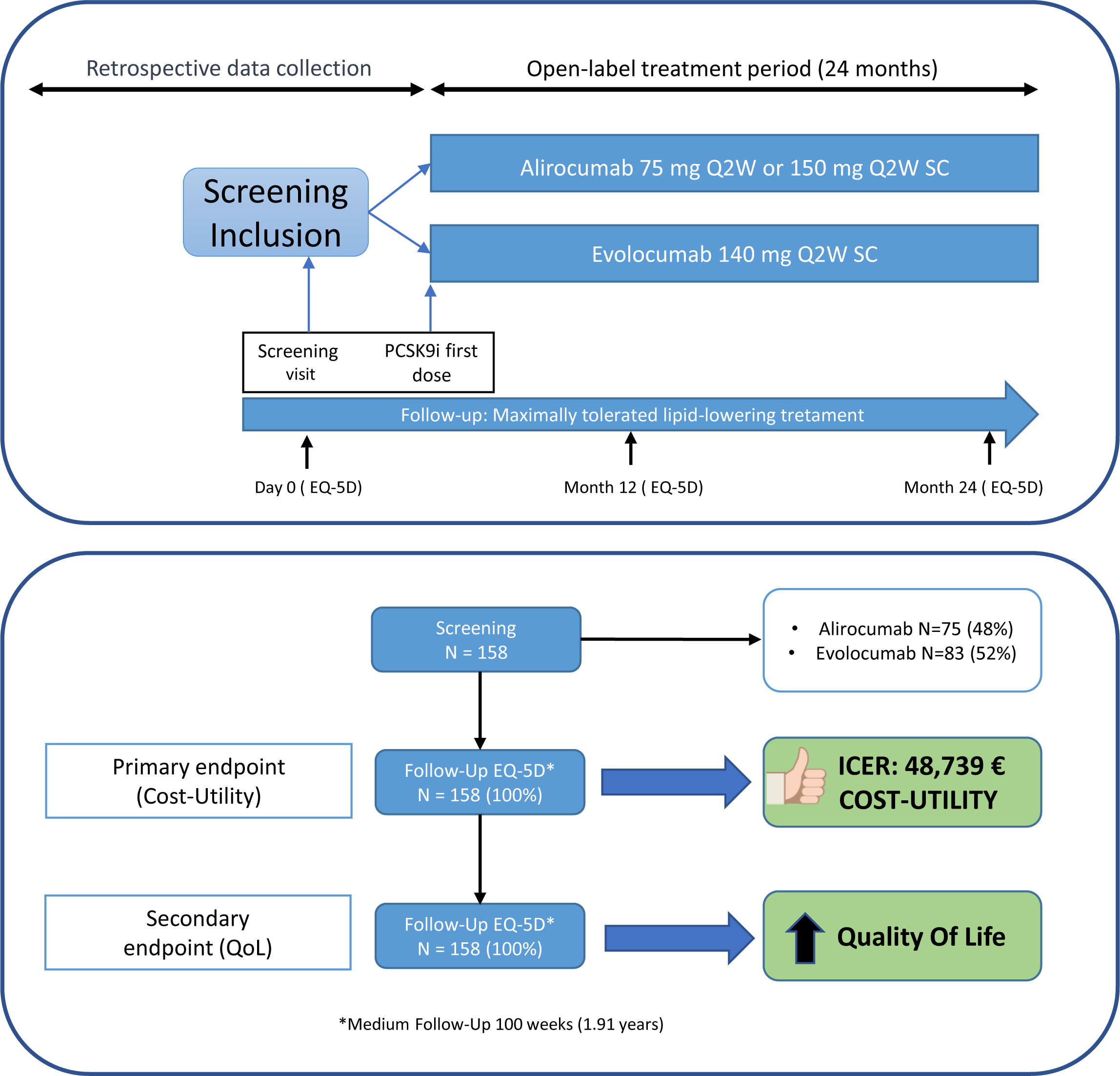

### Population

The inclusion criteria for this sub analysis involved individuals over 18 years old who received their first prescription for a PCSK9 inhibitor: evolocumab (140mg every 2 weeks) or alirocumab (75mg or 150mg every 2 weeks). Eligible participants had established atherosclerotic cardiovascular disease or hypercholesterolemia. Established atherosclerotic cardiovascular disease was defined as having a history of myocardial infarction, stroke, or peripheral arterial disease, while hypercholesterolemia encompassed homozygous familial hypercholesterolemia, heterozygous familial and non-familial hypercholesterolemia, or mixed dyslipidemia.

A total of 158 subjects met the inclusion criteria for this sub analysis and provided written informed consent. The study was conducted in compliance with the principles of the Declaration of Helsinki.

### Study procedures

The inclusion period for this study spanned from May 2020 to May 2022, while the follow-up period extended from May 2020 to February 2023. The EQ-5D-3L questionnaire was administered at baseline, 12 months, 24 months, or during the final visit for patients with a follow-up duration exceeding or falling short of 24 months. The responsibility for questionnaire administration during study visits rested with the investigators, as listed in Appendix 1. Investigators received instructions at the start of the study and during several investigator meetings throughout the follow-up period.

### End points

The primary objective was to conduct a cost-utility analysis over a 2-year period, utilizing the gained quality-adjusted life years (QALYs) and the associated costs of PCSK9 inhibitors compared to basic therapies such as statins or ezetimibe. In this study, the population included for comparison purposes was drawn from the same patient cohort and collected retrospectively, as we assessed the quality of life and costs before the initiation of PCSK9 inhibitor (iPCSK9) treatment and after two years of treatment. Furthermore, cost events and costs of premature deaths were calculated. The Major Adverse Cardiovascular Events (MACE) analyzed were: myocardial infarction, unstable angina, percutaneous coronary intervention, cardiac surgery (Bypass), cardiovascular death and death for any cause.

The secondary objective included evaluating changes in quality of life using the EQ-5D-3L questionnaire. This questionnaire consists of two sections: the EQ-5D descriptive system, which assesses 5 dimensions (MOBILITY, SELF-CARE, USUAL ACTIVITIES, PAIN/DISCOMFORT, and ANXIETY/DEPRESSION) at three levels (no problems, some problems, extreme problems), and the EQ-5D visual analogue scale (VAS) ranging from 0 to 100 points (10). Assessments were conducted at baseline, 12 months, 24 months, and/or at the end of the study. A sample of the EQ-5D-3L is provided in Appendix 2.

### Cost-utility analysis

The cost-utility analysis aimed to assess the economic value of the intervention, primarily using Quality-Adjusted Life Years (QALYs) as the outcome measure. Utility values were derived from the EuroQol Five-Dimensional Questionnaire (EQ-5D), administered at baseline and follow-up assessments. The EQ-5D provides values for all health states, resulting in an index for each of the 243 possible health states, including death, reflecting population preferences (10). Direct healthcare costs, including medication expenses and healthcare resource utilization, were collected from medical records and billing databases (11). Indirect costs, such as productivity loss, were estimated using standard approaches. Cost-effectiveness ratios, expressed as the incremental cost per QALY gained, were calculated to evaluate the efficiency of the intervention compared to alternative treatment strategies.

Regarding the costing perspective, the analysis adopts a societal viewpoint, encompassing both direct healthcare costs and indirect societal costs. Direct healthcare costs were collected from medical records and billing databases. Indirect costs, such as productivity loss, were estimated using standard approaches. Furthermore, discounting was applied to both costs and QALYs at a rate of 3.5%, in line with the standard practice in economic evaluations in healthcare. (12)

To provide a comprehensive overview of the Spanish healthcare system, we should mention that the study was conducted within the framework of the Spanish National Health System, which provides universal coverage to the Spanish population.

In the case of premature death costs, two distinct scenarios were considered:

For the active population: The expected benefits in terms of reduced incidence, mortality, and potential years of lost work-life, estimating the economic value derived from lost wages based on the average gross income per worker in the area (€20,286 per year).

For the working-age population, the cost was estimated taking into account the average gross income of the worker (€18,768.21 per year) and the unemployment rate (9.37%) in our healthcare area as of April 30, 2023 (13). On the other hand, leisure time was valued at 47% of the cost of working hours (14).

For the retired Population: The expected benefits in terms of reduced incidence, mortality, and potential years of life lost (relative to the average life expectancy) were estimated, considering the economic value derived from the contribution of individuals aged 65 and older to volunteer work and grandchild care (IPREM: €600 per month). For the retired population, the cost was estimated taking into account the percentage of individuals aged 65 or older engaged in volunteer work, according to the CIS-IMSERSO study (2.3%) (15); those who dedicate themselves to grandchild care according to the study “Living Conditions of Older People” conducted by the Center for Sociological Research (22.6%) (Center for Sociological Research. Study “Living Conditions of Older People”) (16).

### Statistical analysis

For the primary and secondary endpoints related to EQ-5D-3L domains, statistical analysis was performed using the Fisher test. Also, frequencies and percentages were calculated to describe these domains. To test de VAS scale, mean with their 95% confidence intervals (CI) at and the standard deviation was calculated. Differences were assessed by t-Student test to paired data. Both for the EQ-5D-L3 and for the VAS test, were compared data at the baseline time with the end of follow-up.

Statistical difference was accepted at p<0.05. All analyses were performed using SPSS 19.0 (IBM Corp. Released 2010. IBM SPSS Statistics for Windows, Version 19.0. Armonk, NY: IBM Corp.)

## RESULTS

### Patient disposition

From May 25, 2020 (first patient) to April 6, 2022 (last patient included), a total of 158 patients were enrolled for the study followed for a median of 99 weeks. All participants successfully completed the final EQ-5D questionnaire, except for those who were deceased (n=2), for whom data were collected during previous visits. (Figure 1). Additionally, the entirety of the 158 patients was retrospectively used as a control group before the initiation of iPCSK9 treatment while they were on standard therapy (statins and/or ezetimibe).

### Baseline characteristics and concomitant medication

Table 1 presents the baseline characteristics of the study population. The patients had a mean (SD) age of 61 (10) years, with 66.5% being male. The mean (SD) body weight and BMI were 81 (16) kg and 29 (5) kg/m2, respectively. Among the participants, 85% had cardiovascular ischemic disease (CVD), 25% had familial hypercholesterolemia (FH), 55% had hypertension, 22% had type 2 diabetes (T2D), and 17% had heart failure. Comorbidities included a family history of dementia in 20% of patients, 11% were smokers, and 72% adhered to a diet.

**Table 1.** Demographic, baseline characteristics and treatments

|  |  |
| --- | --- |
| Sex (male); n (%) | 105 (66,5) |
| Years; mean (SD) | 60,6 (10,2) |
| Height; mean (SD) | 1,67 (0,08) |
| Weight; mean (SD) | 81,0 (15,7) |
| Medical history; n (%) |  |
| Cardiovascular disease | 134 (84,8) |
| Familiar hypercholesterolemia | 39 (24,7) |
| Statins intolerance | 69 (43,7) |
| Dementia history | 31 (19,6) |
| Diabetes | 35 (22,2) |
| Hypertension | 87 (55,1) |
| Heart failure | 27 (17,1) |
| Diet | 114 (72,2) |
| Smoking status; n (%) |  |
| Current | 17 (10,8) |
| Past smoker | 85 (53,8) |
| Never | 56 (35,4) |
| PCSK9 inhibitors; n (%) |  |
| Alirocumab 150mg | 65 (41,1) |
| Alirocumab 300mg | 18 (11,4) |
| Evolocumab 240mg | 75 (47,5) |
| Statins; (%) |  |
| Rosuvastatin | 33,5 |
| Atorvastatin | 18,4 |
| Pitavastatin | 3,2 |
| Other statins | 1,2 |
| Ezetimibe | 58,6 |
| LDL-c; mg/dL (SD) |  |
| Baseline | 145,18 (43,43) |
| Follow-up | 62,11 (57,00) |

Out of the 158 patients, 75 were receiving evolocumab 140mg every 2 weeks (47.46%), 65 were on alirocumab 150mg every 2 weeks (41.14%), and 18 were on alirocumab 300mg every 2 weeks (11.40%). As for additional lipid-lowering therapy, 33.5% were taking rosuvastatin, 18.4% were taking atorvastatin, 3.2% were taking pitavastatin, 1.2% were taking other statins, and 58.6% were taking ezetimibe. Notably, 43.7% of the sample was not taking any statin due to statin intolerance.

### Outcomes

#### Primary endpoint: Cost-utility

We conducted an analysis to calculate the gained QALYs and associated costs for both treatments, PCSK9 inhibitors and standard therapy.

The events and costs are showed in table 2 and 3.

**Table 2.** Events.

**Number of de events**
|  | IAM_Nº | Angina<br>Inestable_Nº | stroke_Nº | PCI_Nº | CABG_Nº | Death_cv |
| --- | --- | --- | --- | --- | --- | --- |
| **PCSK9i** | 3 | 2 | 0 | 1 | 0 | 2 |
| **Standard** | 4 | 3 | 1 | 2 | 2 | 0 |

\*Death for other causes were not included into analysis in both arms (2 deaths due to cancer)

**Table 3.**
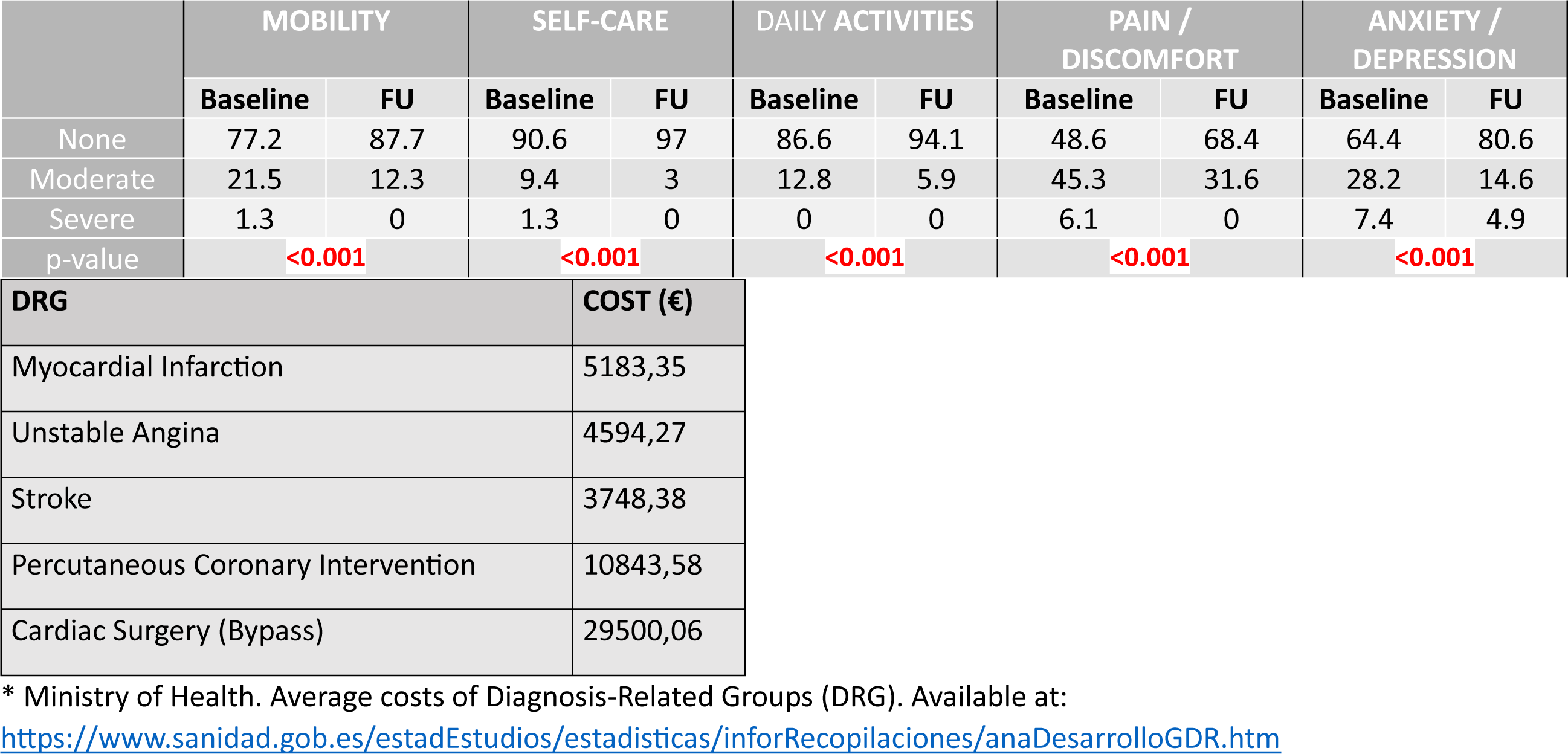
Event costs.

Results:

Pharmacological treatment cost PCSK9 inhibitors: €13,633.39

Pharmacological treatment cost standard therapy: €3,638.25

Total event cost PCSK9 inhibitors: €35,582.17

Unit event cost PCSK9 inhibitors: €483.20092

Total event cost standard therapy: €118,951.87

Unit event cost standard therapy: €1002.35

Total premature death cost PCSK9 inhibitors: €41,246.77

Total premature death cost standard therapy: €40,421.84

Unit cost of events + mortality PCSK9 inhibitors: €483.20

Unit cost of events + mortality standard therapy: €1002.35

Discount rate: 3.5% (16)

Pharmacological treatment cost PCSK9 inhibitors after discount rate: €13,152.56

Pharmacological treatment cost standard therapy after discount rate: €4,323.69

Qualys PCSK9 inhibitors (2 years): 1.648851948

Qualys PCSK9 inhibitors (2 years) after discount rate: 1.53922094

Qualys standard therapy (2 years): 1.454807792

Qualys standard therapy (2 years) after discount rate: 1.35807864

ICER: (Total unit cost PCSK9 inhibitors - Total unit cost standard therapy) /(Qualys PCSK9 inhibitors - Qualys standard therapy) = € 51,427.72

#### Secondary endpoint: Changes in QoL

At the end of the study, there were no missing data for any patients (0%) during the follow-up period. A total of 158 subjects were included in the analysis. Percentage of changes in the EQ-5D domains and the total VAS score were calculated (see table 4 and 5)

**Table 4.** Secondary Endpoint: Changes in Quality of Life Proportions reporting levels within EQ-5D dimensions: Baseline and Follow-Up.

**Proportions reporting levels within EQ-5D dimensions: Baseline and Follow-Up**
|  | MOBILITY |  | SELF-CARE |  | DAILY ACTIVITIES |  | PAIN /<br>DISCOMFORT |  | ANXIETY /<br>DEPRESSION |  |
| --- | --- | --- | --- | --- | --- | --- | --- | --- | --- | --- |
|  | Baseline | FU | Baseline | FU | Baseline | FU | Baseline | FU | Baseline | FU |
| None | 77.2 | 87.7 | 90.6 | 97 | 86.6 | 94.1 | 48.6 | 68.4 | 64.4 | 80.6 |
| Moderate | 21.5 | 12.3 | 9.4 | 3 | 12.8 | 5.9 | 45.3 | 31.6 | 28.2 | 14.6 |
| Severe | 1.3 | 0 | 1.3 | 0 | 0 | 0 | 6.1 | 0 | 7.4 | 4.9 |
| p-value | <0.001 |  | <0.001 |  | <0.001 |  | <0.001 |  | <0.001 |  |

\* % of patients

**Table 5.** Changes in the mean EVA score: Baseline and Follow-Up

|  |  |  |  |  |
| --- | --- | --- | --- | --- |
| EVA | Baseline | 67.04(±20.069) | 66.62 - 72.67 | 0.086 |
|  | End point | 69.64(±18.683) | 63.81 - 70.27 |  |

In the baseline assessment of the Mobility domain, the distribution of symptoms among patients was as follows: 77.2% reported no symptoms, 21.5% had moderate symptoms, and 1.3% experienced severe symptoms. However, at follow-up, the distribution shifted significantly, with 87.7% of patients reporting no symptoms, 12.3% experiencing moderate symptoms, and no patients with severe symptoms. This observed improvement in mobility outcomes was statistically significant (p<0.001).

In the Self-care domain, at baseline, the distribution of patients’ self-care abilities was as follows: 90.6% reported no difficulties, 9.4% had moderate difficulties, and 1.3% experienced severe difficulties. However, at follow-up, there was a significant improvement in self-care outcomes, with 97% of patients reporting no difficulties, 3% experiencing moderate difficulties, and none of the patients reporting severe difficulties.

In the Daily Activities domain, at baseline, the distribution of patients’ abilities to perform daily activities was as follows: 86.6% reported no difficulties, 12.8% had moderate difficulties, and 0.6% experienced severe difficulties. However, at follow-up, there was a significant improvement in daily activities outcomes, with 94.1% of patients reporting no difficulties, 5.9% experiencing moderate difficulties, and none of the patients reporting severe difficulties.

In the baseline assessment of the Pain or Discomfort domain, the distribution of symptoms among patients was as follows: 48.6% reported no pain or discomfort, 45.3% had moderate pain or discomfort, and 6.1% experienced severe pain or discomfort. However, at follow-up, the distribution shifted significantly, with 68.4% of patients reporting no pain or discomfort, 31.6% experiencing moderate pain or discomfort, and no patients with severe pain or discomfort. This observed improvement in pain or discomfort outcomes was statistically significant (p<0.001).

Finally, regarding the baseline assessment of Anxiety or Depression domain, the distribution of symptoms among patients was as follows: 64.4% reported no anxiety or depression, 28.2% had moderate levels of anxiety or depression, and 7.4% experienced severe levels. However, at follow-up, the distribution shifted significantly, with 80.6% of patients reporting no anxiety or depression, 14.6% experiencing moderate levels, and 4.9% with severe levels. This observed improvement in anxiety or depression outcomes was statistically significant (p<0.001).

The mean change in VAS score from baseline was 67.04 (±20.069) (95% CI 66.62-72.67) to follow-up, with a value of 69.64 (±18.683) (95% CI 69.64-18.683) (p=0.086). This represents an increase of 2.6 points, although the difference was not statistically significant.

See figure 2.

**Figure 2.**
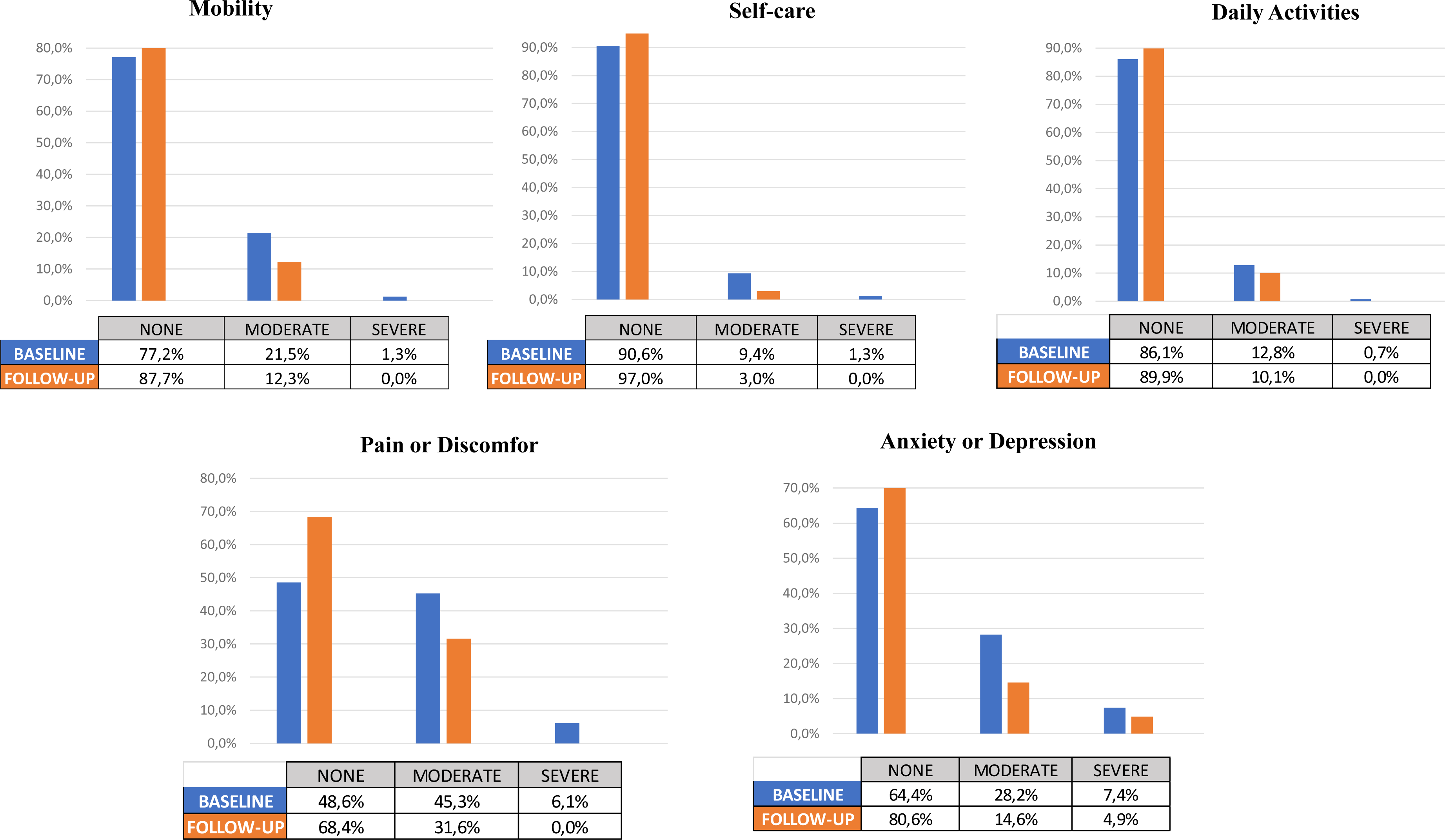

## DISCUSSION

The analyses conducted in this observational and prospective study, which included real-world patients treated with PCSK9 inhibitors, provide valuable insights into the cost-utility of PCSK9 inhibitors in this population. The results of the study demonstrate that the use of PCSK9 inhibitors was associated with favourable cost-utility outcomes, indicating its potential economic value in real-world clinical practice. Additionally, we also assessed changes in quality of life using the EQ-5D-3L questionnaire among 158 real-world patients followed from the initiation of treatment with PCSK9 inhibitors for a duration of 2 years. The results obtained indicate an improvement in all domains: mobility, self-care, activities, pain, and anxiety and/or depression. Furthermore, there was an improvement observed in the overall VAS EQ-5D-3L score.

Among the 158 patients included in this study, the groups taking alirocumab and evolocumab were were almost equally distributed, and 158 patients completed the EQ-5D questionnaire at follow-up (100%) (figure 1). The main reduction in LDL-c levels was 55.6% (from 145.18 mg/dl to 62.11 mg/dL), which closely aligns with the results reported in the pivotal RCT (3,4). Major Adverse Cardiovascular Events (MACE) were observed in 10 patients following the initiation of PCSK9 inhibitors during a 2-year follow-up period. As depicted in Table 2, we also retrospectively recorded events related to standard therapy from the electronic medical record system (IANUS), which covers all hospitals and primary care centers in this area. All direct and indirect costs were obtained from the Ministry of Health, and the average costs of Diagnosis-Related Groups (DRG) (16) were utilized, with appropriate discounts and updates applied.

In relation to the primary outcome, the calculated incremental cost-effectiveness ratio (ICER) was €48,739.97. This value suggests that the use of PCSK9 inhibitors demonstrates favourable cost-utility across various scenarios and when compared to different comparative references. The ICER serves as an important indicator of the efficiency and economic value of an intervention, and the calculated value supports the cost-effectiveness of utilizing PCSK9 inhibitors in the context of this study. These findings highlight the potential benefits and value of incorporating PCSK9 inhibitors into clinical practice, considering their impact on both cost and utility in comparison to alternative therapies.

One of the economic scientific comparators, A. Laupacis et al. (17), indicates a threshold of €51,278.21 ($55,000 US), which, when updated to January 2023, would be € 94,076.4. This suggests that the intervention clearly demonstrates cost-utility. When referring to Plans P. et al as a comparative reference (18), the results also indicate cost-utility, as the economic threshold for this comparator is €46,616,562 ($50,000 US). If we update it to January 2023, the ICER limit would be € 68,420. Considering the De Cock & González-Juanatey study (19), which indicates a threshold range between €12,000 and €45,000, even with the current updated value of € 52,965 for January 2023, the study still demonstrates cost-utility. Finally, the scenario suggested by the CHOICE project of the World Health Organization (WHO) (20) considers a drug to be cost-effective if its cost-utility is between 1 and 3 times the Gross Domestic Product (GDP) per capita. In Spain, the GDP per capita in 2022 was €27,820, which aligns with the results of this study.

Regarding changes in EQ-5D, it is evident that significant improvements were observed in all domains of the EuroQol-5 Dimension (EQ-5D) questionnaire. Furthermore, when comparing the baseline and follow-up scores, the domains with the most pronounced improvements were Pain/Discomfort and Anxiety/Depression. It is important to note that although improvements were observed across all domains, the Pain/Discomfort and Anxiety/Depression domains showed the most remarkable changes. These findings suggest that the use of PCSK9 inhibitors may have a particularly beneficial effect on reducing pain, discomfort, anxiety, and depression among the study population. Our findings align with the direction observed in clinical trials regarding the positive effects of PCSK9 inhibitors on quality of life. For instance, the FOURIER and ODYSSEY trials (3,4) reported improvements in health-related quality of life measures among patients treated with PCSK9 inhibitors compared to standard therapy.

To the best of our knowledge, this is the first real-world cost-utility study to date. Most of the existing studies have focused on cost-effectiveness rather than cost-utility. For example, Samad Azari et al. (21) conducted a cost-effectiveness systematic review comparing PCSK9 inhibitors with standard therapy but do not directly align with our cost-utility analysis. It is important to note that our study results may differ from those of clinical trials. One possible explanation is that patients in our real-world study, treated with iPCSK9 inhibitors, may have experienced fewer events compared to patients in clinical trials. This difference could be attributed to various factors, including differences in patient characteristics or comorbidities, and treatment adherence. Additionally, the duration of follow-up in our study may have been shorter than that of clinical trials, which could influence the occurrence of events. On the other hand, several published studies (22–25) were in line with our study, and they supported to be cost-effectiveness but none of them studied the cost-utility. Based on these considerations, we believe that this study provides a novel economic analysis on the real-world impact of iPCSK9 inhibitors. However, further comparative studies in this regard will be necessary to strengthen the evidence base. These additional studies can contribute to a more comprehensive understanding of the cost-utility profile of PCSK9 inhibitors and help guide decision-making in clinical practice and healthcare policy.

Further research and longer-term follow-up are needed to fully understand the comparative outcomes and economic implications of PCSK9 inhibitors in real-world settings.

### Limitations

Despite the valuable insights provided by our study, there are certain limitations that should be acknowledged. First, as an observational study, it is subject to inherent biases and confounding factors. Although efforts were made to minimize these biases through robust data collection and statistical adjustments, the potential for residual confounding remains.

Second, the study relied on retrospective data collection for certain variables, such as events and costs associated with standard therapy. While efforts were made to obtain accurate and comprehensive data from electronic medical records, the completeness and accuracy of these records could vary, potentially impacting the results. Additionally, it’s worth noting that this study has specific limitations as the control group and the treatment group consist of the same patients. However, this potential bias has been mitigated by the fact that data collection spans two years prior to the initiation of PCSK9i treatment, aligning with the follow-up period for the treatment under investigation.

Another limitation is the relatively short follow-up period of 2 years. Longer-term follow-up would provide a more comprehensive understanding of the cost-utility outcomes and potential changes in health-related quality of life over time.

Finally, it is important to note that cost-utility analyses rely on certain assumptions and models to estimate the economic outcomes. These assumptions may introduce uncertainty and limit the generalizability of the results to different healthcare systems or contexts.

## CONCLUSIONS

The current study represents a significant contribution to the literature by being one of the first real-world cost-utility analyses conducted on the use of PCSK9 inhibitors. Our results not only demonstrate favorable cost-utility outcomes but also reveal an improvement in all domains of the EQ-5D questionnaire. This suggests that the incorporation of PCSK9 inhibitors into routine clinical practice is not only economically justified but also holds the potential to enhance patients’ quality of life. These findings provide valuable insights for healthcare decision-makers along with reimbursement policies, as they highlight the potential benefits of PCSK9 inhibitors in terms of both cost and utility.

### Statements & Declarations

#### Funding

The authors declare that no funds, grants, or other support were received during the preparation of this manuscript.

#### Conflict of interest

The authors have no relevant financial or non-financial interests to disclose.

#### Availability of data and material (data transparency)

In this article, we are committed to data transparency and availability of materials used in our research. The data and materials will be available on demand for readers interested in replicating or building upon our study. Interested parties can contact the corresponding author to request access to the data and materials. We will ensure that the requested information and materials are provided in a timely and complete manner.

#### Code availability (software application or custom code)

Not applicable

#### Authors ‘contributions

All authors contributed to the study conception and design. Material preparation, data collection and analysis were performed by Jose Seijas-Amigo, Ana Estany and Pedro Suarez-Artime. The first draft of the manuscript was written by Jose Seijas-Amigo and all authors commented on previous versions of the manuscript. All authors read and approved the final manuscript. All named authors meet the International Committee of Medical Journal Editors (ICMJE) criteria for authorship for this article.

#### Ethics approval

This study was performed in line with the principles of the Declaration of Helsinki. Approval was granted by the Ethics Committee of Servizo Galego de Saúde (Date 29-JAN-2020 /No. 2019/653) and Agencia Española de Medicamentos y Productos Santiarios (Date 28-JAN-2020)

#### Consent to participate

Informed consent was obtained from all individual participants included in the study. Informed consent was approved by the Ethics Committee of Servizo Galego de Saúde (Date 29-JAN-2020/No. 2019/653).

#### Consent to publish

Not applicable

## Data Availability

Availability of data and material (data transparency) In this article, we are committed to data transparency and availability of materials used in our research. The data and materials will be available on demand for readers interested in replicating or building upon our study. Interested parties can contact the corresponding author to request access to the data and materials. We will ensure that the requested information and materials are provided in a timely and complete manner.

## Acknowledgments

Investigators received the support of the Fundación Instituto de Investigación Sanitaria de Santiago de Compostela (FIDIS) and National Network for Biomedical Cardiovascular Research of Cardiovascular Disease (CIBERCV, Centro de Investigación Biomédica en Red de Enfermedades Cardiovasculares). Bonnie Dyer contributed to the English writing assistance.

## Notes

### Competing Interest Statement

The authors have declared no competing interest.

### Clinical Trial

Trial Registration clinicaltrials.gov Identifier: NCT04319081

### Author Declarations

Ethics approval This study was performed in line with the principles of the Declaration of Helsinki. Approval was granted by the Ethics Committee of Servizo Galego de Saúde (Date 29-JAN-2020 /No. 2019/653) and Agencia Española de Medicamentos y Productos Santiarios (Date 28-JAN-2020)

